# A large dataset of brain imaging linked to health systems data: the curation and access to a whole system national cohort from NHS Scotland

**DOI:** 10.1101/2025.10.21.25338469

**Authors:** Michael P J Camilleri, Dorian Gouzou, Salim Al-Wasity, Muthu R K Mookiah, María Valdes Hernandez, Bea Alex, Sotirios A. Tsaftaris, Andrew Brooks, Ruairidh MacLeod, Honghan Wu, Brenda Bauer, Claire Grover, Parminder Reel, Susan Krueger, Richard Tobin, J. Douglas Steele, Grant Mair, Joanna Wardlaw, Alexander Doney, Emanuele Trucco, William Whiteley

**Author notes:** equal contribution (Michael P J Camilleri, Dorian Gouzou). equal contribution (Emanuele Trucco, William Whiteley).

## Abstract

We present the design and implementation of a data curation framework to generate a large-scale clinical brain imaging dataset suitable for artificial intelligence (AI) enabled image analysis, accessible through the Brain Health Data (BHD) initiative. The raw data accessible through the BHD includes approximately 417K magnetic resonance imaging (MRI) and 846K computerized tomography (CT) head scans, linked electronic health records (EHRs), and associated free-text imaging reports from clinical practice between 2010 and 2018 in Scotland, totally exceeding 185 TB storage of brain imaging and associated data.

We present the work curating the dataset and the strengths of the BHD, including clinical relevance thanks to its unprecedented scale, population-wide representativeness of a national free-at-the-point-of-service healthcare, long-term follow-up to neurodegenerative disease, and real-world variability. We discuss challenges and lessons learnt in developing the framework to curate the data initially available, including the time needed to obtain relevant permissions, the need for easily accessible, secure, responsive and affordable computational environments, the variability and inconsistencies of clinical data and records, and the challenge of extracting linked clinical data and images at scale, among others. This resource will be crucial for clinical research, fostering the development of personalized medicine approaches, and fast-tracking the implementation of AI models in clinical workflows. We encourage the use of the BHD data through a streamlined application to the Data Research and Innovation Service (eDRIS) of Public Health Scotland (PHS).

## INTRODUCTION

Brain imaging plays a crucial role in the diagnosis of neurological disorders. However, clinical imaging services are under great demand, highlighting the need for new tools to improve radiology workflows. These tools should accelerate image assessment, reduce the workload for radiologists, and ultimately improve patient care. Artificial intelligence (AI) methods show promise for faster diagnosis (for example in acute ischaemic stroke)[1] and similar improvements are possible in head injury, neurodegeneration and brain cancers[2,3] To develop and test AI models that are clinically relevant, researchers need access to large datasets of clinically acquired images, with sufficient computing resources, and secure, ethical data provision.

We present the Brain Health Data (BHD) initiative, a framework that unifies all clinical brain imaging data with related information collected in Scotland, to facilitate their access through the Electronic Data Research and Innovation Service (eDRIS) of Public Health Scotland (PHS) for other researchers. The BHD dataset includes magnetic resonance imaging (MRI) and computerized tomography (CT) head scans, linked electronic health records (EHRs), and free text radiology reports. We also present the design and implementation of the MRI data processing framework to curate this large-scale clinical brain imaging dataset in a format suitable for AI analysis, which we make available through the BHD. The work was undertaken during the SCANDAN project (SCottish AI in Neuroimaging to predict Dementia and Neurodegenerative Disease), which aimed to develop AI algorithms for reliable dementia risk estimation with routine brain imaging and clinical records. The SCANDAN work packages are illustrated in Figure 1. SCANDAN used Scotland-wide clinical practice data from 2010 to 2018 from more than 830 thousand individuals, now made accessible through the BHD. The BHD dataset offers clinical relevance with its unprecedented scale, population-wide representativeness of healthcare, long-term follow-up to neurodegenerative disease, and real-world variability.

**Figure 1.**
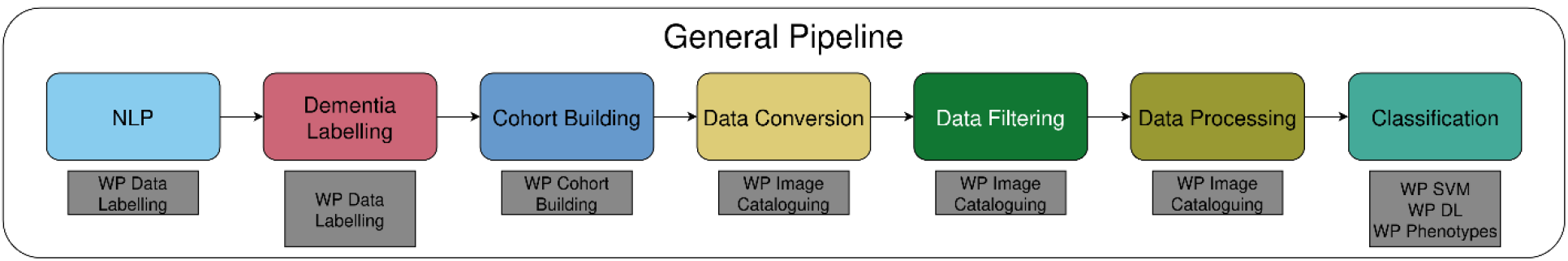
Work packages (WP) in the SCANDAN project: Data labelling, cohort building, image cataloguing, and processing for classification into being indicative of having dementia or not using deep learning (DL), support vector machine (SVM) and from the analysis of extracted imaging phenotypes.

A survey conducted between December 2024 and February 2025 across UK secure data environments found no comparable brain imaging resource with nationwide coverage. For instance, the Diagnostic Imaging Dataset curated by NHS England includes patient-level metadata on the 501 million diagnostic imaging procedures performed within NHS England facilities since April 2012, but it lacks actual imaging data and associated reports.[4]

The preparation of large repositories of routinely collected imaging data is challenging, particularly within secure data environments that are designed to protect patient privacy. Despite adherence to Digital Imaging and Communications in Medicine (DICOM)[5] standards as best practice, real-world medical imaging datasets vary significantly in quality, format, and acquisition protocols, which makes standardization across different imaging sources necessary. Determining imaging sequences (e.g., T1-or T2-weighted MRI) is essential for analysis, but difficult in practice. DICOM meta-data tags provide rapid but sometimes unreliable classification, while image-based classification is more accurate but computationally demanding, and not free from ambiguities. Natural language processing (NLP) of radiology reports can aid in sequence identification if recorded in the free text.

Filtering out unusable scans, such as those with artefacts or missing brain structures, is a critical step. Automated AI-based image quality assessment can help, but manual verification may be necessary. The automation of data pipelines, and the integration of structured clinical records with imaging data, can facilitate data retrieval and pre-processing. Compliance with governance frameworks is important to access large-scale unconsented clinical imaging datasets within safe havens, and needs ethical approval, data governance agreements, and compliance with privacy regulations, all with costs. These administrative barriers, although necessary, can significantly delay or completely deter research. To address these challenges, SCANDAN developed an automated framework that extracted, cleaned and prepared structured brain imaging and clinical data for AI analysis, creating a research-ready dataset. This paper presents SCANDAN’s methods, highlights key lessons learned from working within Scotland’s data governance frameworks, and describes how to access the data through the BHD.

## METHODS

### Data sources

PHS’ eDRIS provided brain CT and MR head studies in adults performed in Scotland between 2010 and 2018 from the Scottish Medical Imaging (SMI) service [6]. S*tudy* refers here to a complete imaging session, encompassing all images obtained during a single scanning session. Each scan contains three hierarchical levels: study, series, and images. Within each study, there are one or more series that group together images acquired using the same imaging technique or settings. Each series is, in turn, composed of multiple single images or “slices”. We linked each study (i.e., one per person) deterministically with pseudonymized identifiers based on the Community Health Index (CHI) number which is the unique patient identifier used across NHS Scotland. We linked them with hospital admission records (SMR01), dementia records from mental health hospitalisations (SMR04), dispensed prescriptions from Prescribing Information System (PIS), death records (National Records of Scotland (NRS)) and demographics (birth year, sex, deprivation index) since the year 2000. All the data was processed and stored within the National Safe Haven (NSH) a secure data environment provisioned by the Edinburgh Parallel Computing Centre (EPCC) (https://edinburgh-international-data-facility.ed.ac.uk/services/safe-haven-services/scottish-national-safe-haven).

### Permissions and research governance

For SCANDAN, multicentre research ethics permission from the NHS Human Research Authority was obtained from the North of Scotland Research Ethics Committee (23/NS/0017). Permission to access the data was provided by the NHS Scotland Public Benefit and Privacy Panel for Health and Social Care (HSC-PBPP), a patient advocacy panel which scrutinises applications for access to NHS Scotland health data for non-direct care (PBPP application 2223-0200 and 2223-0005). The SCANDAN team engaged with several Scottish public and patient groups during the application process and throughout the project.

### Computing environment

The Scottish NSH is hosted in EPCC’s Trusted Research Environment (TRE), a secure infrastructure which currently hosts twelve Safe Havens. Each Safe Haven is operated under the “Five Safes” framework,[7] designed by the Office for National Statistics, and the Scottish Government Charter for Safe Havens.[8] Researchers access a secure data sharing and analysis environment with a virtual desktop, under the terms and conditions prescribed by the data providers. Standard software packages such as R and Python are available in the NSH; additional software packages can be installed from repositories such as the comprehensive R archive network (CRAN) and the Python package index (PyPI). Safe Havens have access to large shared-memory, high-performance computer clusters, including one with graphical processing unit (GPU) accelerators for large-scale analysis. All EPCC Safe Haven Services are operated at EPCC’s Advanced Computing Facility, located in Edinburgh, Scotland. The EPCC Trusted Research Environment (TRE) is accredited by ISO27001 [9] for information security practices and self-certified under Cyber Essentials and NHS Digital’s Data Security and Protection Toolkit (DSPT). In addition, the NSH is accredited under the Digital Economy Act 2017 by the UK Statistics Authority, and all Safe Havens in the TRE are operated to the same standard.

### Natural language processing of brain imaging reports

We applied a clinical NLP tool, the Edinburgh Information Extraction for Radiology (EdIE-R), [10,11] which was originally developed and validated for radiology reports of brain imaging in the Edinburgh Stroke Study and NHS Tayside [12] EdIE-R processes radiology reports through a pipeline architecture that identifies entities, detects negation, extracts relationships and assigns document-level labels to identify phenotypes referring to brain abnormalities. The tool was later adapted and validated for use with data from other areas in Scotland provided by Generation Scotland.[13] EdIE-R was used to extract 24 distinct phenotypes, including different types of strokes (ischaemic, haemorrhagic and underspecified, with time and location details), brain tumours (meningiomas, gliomas, metastases or underspecified), small vessel disease, microbleeds, atrophy and other neurological abnormalities. Additionally, it marked up MRI sequence types (T1, T2, and FLAIR) to support further analysis.

To improve data selection, EdIE-R was enhanced to identify scans of non-head and non-brain body parts, and flag them for exclusion. This ensured that only relevant imaging reports were included in the analysis. Enhancement of the detection of section boundaries allowed more precise separation of clinical history from the main report text so that phenotype mentions were only extracted from the latter. The refined EdIE-R pipeline was applied to all radiology reports in the SCANDAN project, producing structured outputs to guide data selection for image analysis and predictive modelling. By processing radiology reports within the Scottish NSH, the tool allows exclusion of scans (e.g., those showing tumours or non-brain regions) and served to validate outputs from imaging type classification and phenotype extraction. This process streamlined the subsequent image analysis and predictive modelling processes.

### Phenotyping dementia

We defined dementia with ICD-10 codes or prescribed medicines that are generally used for Alzheimer’s disease (AD) [14]. Each patient interaction with the health system was used, including a single stay in hospital, multiple consecutive stays, a prescription, or a death record. We defined labels for ‘any dementia’ and five dementia subtypes: AD, vascular dementia, other rare dementias, unspecified dementia and possible dementia. The subtype was defined as the most frequently occurring dementia phenotype in each person’s electronic record. (**Error! Reference source not found**.). All individuals with a dementia label were categorised as cases, and individuals with no mention of dementia in any record were considered controls.

### Cohort building

To test the BHD data capability, we built a matched case-control study cohort with MR brain images. A matched case-control design was chosen for several reasons. First, most deep learning and other algorithms work best with balanced cases and controls. Second, we had limited computing capacity at the beginning of the project. Third, the rate of image delivery was limited by the need to copy data from a preparation area to a research area which had limited storage capacity. We first selected useable images from those that had gone through the data pipeline to define image sequence, ensure presence of whole brain, absence of other body parts and absence of intravenous contrast, and had no NLP label of tumour or haemorrhagic stroke in the radiology report. We selected people who were aged over 40 years at the time of scan or had an associated EHR record. To exclude people with a dementia diagnosis at time of scan, we defined controls as those without a diagnosis of dementia at any point in their record, and cases where a diagnosis of dementia was made one year or more after their first scan.

Dementia cases were matched to controls based on age at the time of scan (within one year of the matched case) and recorded sex from the linked demographic information. Cases with no matched controls were discarded. Age and sex matching was verified by analysing the resulting distributions over the entire cohort.

### Identification of MR scan type and sequence

DICOM tags were used to produce five labels for each image series: imaging sequence, presence of brain, presence of other body part, angiography, and imaging with contrast. We aimed to retain MRI series with sequences T1, T2 and Fluid-Attenuated Inversion Recovery (FLAIR), containing a brain and no other body part than the neck, without angiography or contrast, and with a 3D image volume of over 5 litres. These labels were subsequently combined with the results from the NLP tool, the MRI acquisition parameters, and the computed volume of the image series, to exclude those which did not meet SCANDAN criteria.

To produce these labels, the DICOM tags were parsed with regular expressions. For example, the expressions /(?i)(?<!pa)t2/ (case insensitive and ignoring occurrences starting with “pa”) and /*se2d1/ were associated with the intermediate label “tmp-T2” (T2-weighted), while the expressions /TOF/ and /MRA/ were associated with the intermediate label “tmp-MRA” (MR angiography). Then, the final labels were created by grouping all the intermediate labels of a series. For example, for an image series to be labelled “T1”, it had to match the intermediate label “tmp-T1”, and could optionally match the intermediate labels “FLAIR”, “GRE” (gradient echo), and “FAT SAT” (fat saturation), which are not T1-weighted exclusive, but not any other intermediate label.

For sequence identification, the DICOM tag ‘Series Description’ (0008,103E) was used. To identify the body part, we used the tags ‘Body Part Examined’ (0018,0015), ‘Protocol Name’ (0018,1030), ‘Performed Procedure Step Description’ (0040,0254) and ‘Study Description’ (0008,1030). Angiograms were identified with the tags ‘Angio Flag’ (0018, 00), ‘Study Description’ (0008,1030), ‘Protocol Name’ (0018,1030) and ‘Series Description’ (0008,103E). Contrast identification used ‘Study Description’ (0008,1030), ‘Contrast/Bolus Agent’ (0018,0010), ‘Contrast/Bolus Route’ (0018,1040), ‘Performed Procedure Step Description’ (0040,0254) and ‘Series Description’ (0008,103E).

The results from the NLP tool provided additional information for the sequence identification and the presence of other body parts. Finally, the MRI sequence was defined with the tags ‘Echo Time’ (0018,0081), ‘Inversion Time’ (0018,0082), ‘Repetition Time’ (0018,0080), ‘Scanning Sequence’ (0018,0020), ‘Flip Angle’ (0018,1314), ‘Sequence Name’ (0018,0024) based on “optimal” value [7-8] adapted to the data through observation on manually annotated data, to complement the identification based on the series description.

### Data validation and quality control

To generate ground truth labels for an initial evaluation of the automatic labelling process described above, we developed a custom python-based graphical user interface (GUI) optimized for MRI and CT DICOM files. The GUI allowed users to load DICOM images from a single or nested folder structure. It utilized DICOM header metadata to: stack slices according to the acquisition order using the DICOM tag ‘Instance Number’ (0020,0013), in ascending, descending, or interleave format, constructing and saving the 3D volumes for subsequent analysis; filter CT scans using the tag ‘Modality’ (0008,0060) and adjust their intensities, e.g. brain-windowing, using the tag ‘Rescale Intercept’ (0028,1052); determine the orientation of the imaging planes (i.e., axial, sagittal, or coronal) using the ta’Image Orientation (Patient)’ (0020,0037) to display mid-axial, mid-coronal, and mid-sagittal views for assessment; and 4c alculate the aspect ratio using the ‘Slice Thickness’ (0018,0050) and ‘Pixel Spacing’ (0028,0030) tags for accurate scaling and visualising of mid-view slices within the designated display area.

Five experts (3 clinicians and 2 trained imaging scientists) annotated 713 randomly selected scans (388 MRI, 319 CT) with modality, sequence, presence of contrast, lesion and artefacts, presence of full brain, and presence of body parts (Figure 2). The 713 scans were evenly divided among the five annotators, with a subset of 100 images overlapping for cross-validation. The Generative model of Labels, Abilities, and Difficulties (GLAD) [14] probabilistic framework was used to estimate the true label for each image while accounting for annotator expertise and image difficulty.

**Figure 2.**
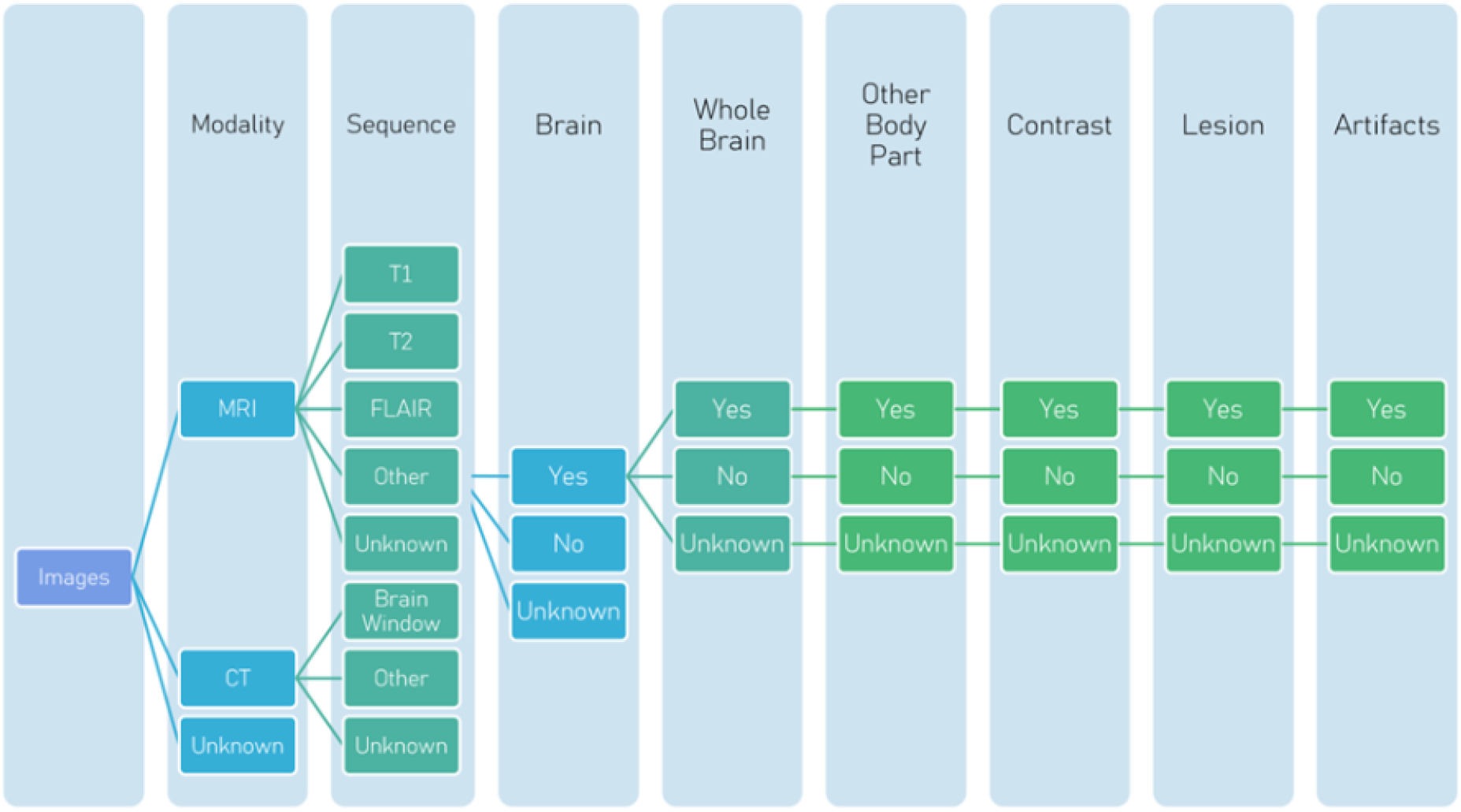
Criteria used by the annotators to label the test imaging set using the GUI developed.

The ground-truth labels were compared with those from the automatic labelling using GLAD. Scans were further reviewed by a neuroradiologist and an experienced imaging scientist independently where the ground truth and automatic labels disagreed regarding sequence type and contrast, or the ground-truth labels were “unknown” or “uncertain”. A third round of annotations resolved disagreements between the neuroradiologist and the imaging scientists regarding the sequence type. The presence of brain, of other body part and the fullness of the brain was re-annotated by a trained image scientist, due to the simpler amount of ‘unknown’ result.

## RESULTS

### Dataset description

MR and CT brain images were available for 830,884 people. Exclusion criteria at subject levels and their effects are listed in **Table *3***. After applying exclusion criteria, 10,709 MRI and 57,242 CT dementia cases were age and sex matched to the same number of healthy controls. The earliest 21,418 MRI and 114,484 CT studies associated with these subjects were requested, of which 21,197 and 94,740 were successfully retrieved.

**Table 1.** International classification of diseases 10 (ICD*-*10) and British national formulary (BNF) codes to define dementia subtypes.

|  | **ICD10** | **BNF** |
| --- | --- | --- |
| Alzheimer's disease | F00* G30* | 0411000 |
| Vascular dementia | F01* |  |
| Other rare dementias | F02*, G31.0, A81.0 |  |
| Unspecified dementia | F03, F05.1 |  |
| Possible dementia | F05.0, G31 |  |

\*Indicates wildcard

**Table 2.**
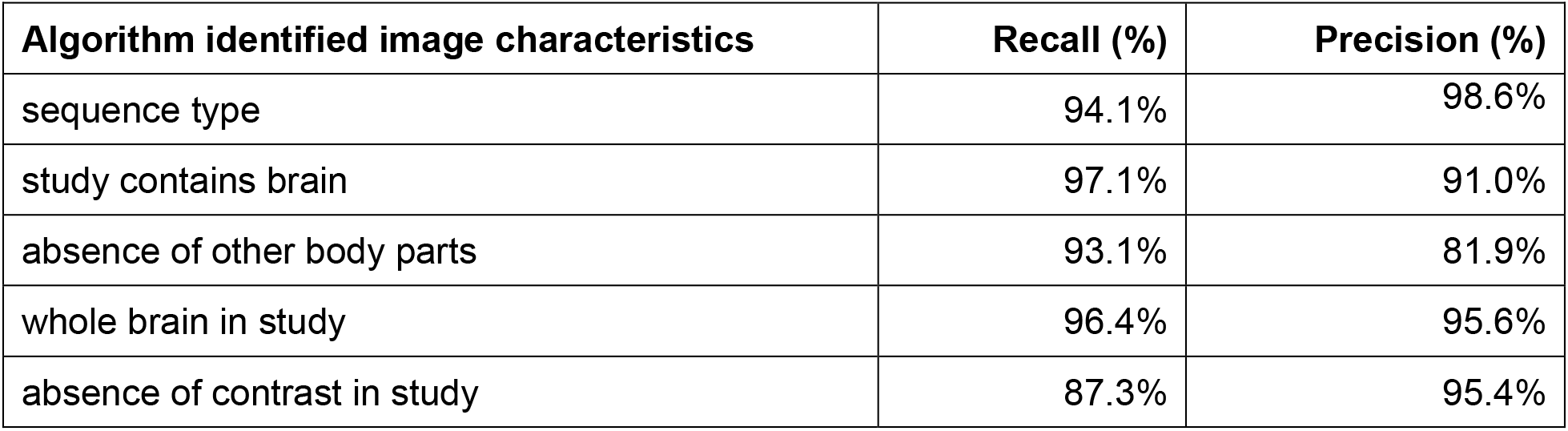
Labels from automated pipeline versus ground truth manual annotation for image characteristics.

| **Algorithm identified image characteristics** | **Recall (%)** | **Precision (%)** |
| --- | --- | --- |
| sequence type | 94.1% | 98.6% |
| study contains brain | 97.1% | 91.0% |
| absence of other body parts | 93.1% | 81.9% |
| whole brain in study | 96.4% | 95.6% |
| absence of contrast in study | 87.3% | 95.4% |

**Table 3.** Selection process for the MRI and CT cohorts, showing both the overall and dementia counts, with addition of characteristics.

| Criterion | MRI |  | CT |  |
| --- | --- | --- | --- | --- |
|  | Overall | Dementia | Overall | Dementia |
| **With scan** | 294,422 | 16,819 | 669,539 | 119,423 |
| + No reported tumour or haemorrhagic stroke | 280,549 | 15,648 | 640,400 | 116,121 |
| + Hospitalised electronic health record | 279,004 | 15,648 | 638,680 | 116,121 |
| + Diagnosis other than 'possible' dementia | 275,970 | 15,648 | 627,911 | 116,121 |
| + Age at scan > 40 | 207,876 | 15,578 | 523,855 | 116,021 |
| + Dementia diagnosis or follow-up > 1 year | 190,582 | 10,709 | 391,356 | 57,242 |

MRI: magnetic resonance imaging of brain; CT: computerised tomography of brain

The MRI cohort contained 8,145 cases (53.2% female) and 7,236 controls (54.1% female) (Table 4). The mean age at scan was 74 years. The mean time from scan to first mention of dementia was 5 years for cases, and the mean follow-up time for controls was 6 years 9 months. Of the 8,145 dementia cases there was non-exclusive record of AD in 3,774, vascular dementia in 3,386, unspecified dementia in 3,784 and other dementia types in 508. The mean number of hospitalisations in the year prior to scan was 1.1 (standard deviation ([SD] 1.52) for cases and 1.0 (SD 1.50) for controls. During the same period, the mean number of prescriptions was 15.4 for cases and 14.2 for controls.

**Table 4.** Distribution of subjects with mention of each dementia type and, controls grouped by key characteristics.

|  |  | **Alzheimer's** | **Vascular** | **Other or rare** | **Unspecified** | **Controls** |
| --- | --- | --- | --- | --- | --- | --- |
| **Sex** | Female | 2,057 | 1,681 | 207 | 2,047 | 3,917 |
|  | Male | 1,717 | 1,705 | 301 | 1,737 | 3,319 |
| **Age in years** | **Mean (SD)** | 73 (8.8) | 75 (8.7) | 67 (9.3) | 74 (8.9) | 74 (9.0) |
|  | 40-50 | 36 | 32 | 19 | 38 | 87 |
|  | 51-60 | 282 | 169 | 90 | 227 | 434 |
|  | 61-70 | 968 | 706 | 179 | 813 | 1611 |
|  | 71-80 | 1,694 | 1,503 | 182 | 1,664 | 3,182 |
|  | 81+ | 794 | 976 | 38 | 1,042 | 1,922 |
| **SIMD** | **Mean (SD)** | 2.8 (1.3) | 2.7 (1.3) | 2.8 (1.3) | 2.7 (1.3) | 2.8 (1.3) |
|  | 1 | 719 | 768 | 102 | 842 | 1429 |
|  | 2 | 732 | 650 | 102 | 724 | 1380 |
|  | 3 | 753 | 635 | 94 | 703 | 1381 |
|  | 4 | 909 | 691 | 113 | 780 | 1523 |
|  | 5 | 363 | 242 | 42 | 325 | 804 |
| **Hospitalisation 1 yr before scan, Mean (SD)** |  | 0.9 (1.4) | 1.2 (1.5) | 1.0 (1.6) | 1.1 (1.5) | 1.0 (1.5) |
| **Prescriptions 1 yr before scan, Mean (SD)** |  | 14.5 (9.5) | 16.1 (0.2) | 14.3 (9.7) | 15.6 (10.2) | 14.2 (9.4) |

SD: standard deviation; SIMD: Scottish index of multiple deprivation

**Table 5.** Final population.

| Sequence | Original | Brain | Whole Brain | Other Body Parts | Contrast & Angio | One Series per Study | Final |
| --- | --- | --- | --- | --- | --- | --- | --- |
| FLAIR | 16,871 | -60 | -211 | -1,742 | -628 | -714 | 13,516 |
| T1 | 27,627 | -1,290 | -4,670 | -2,707 | -3,153 | -2,156 | 13,651 |
| T2 | 28,959 | -1,407 | -4,828 | -3,738 | -890 | -3,297 | 14,799 |

### Data validation and quality control

For simplicity, we refer to the results of the manual annotations as “annotations”, and the results of the automatic tools which are compared to the manual annotations as “labels”.

During the first round, 707 annotations were obtained from 713 images. Four images could not be read due to acquisition errors. Two images were only partially annotated due to visual perception error and discarded. 29 (4.5%) images were wrongly annotated because several scans containing only one slice (localiser) or did not containing a brain. As the annotators were less familiar with CT, 24 (7.52%) of CT images were written as ‘Unknown’. In the labelling process we used the previously validated DICOM tag ‘Modality’ (0008,0060), to identify CT and MRI scans. The identification of the body parts was easier for the annotators than the modality or sequence type. “Unknown” was given for 23 (3.25%) series when annotators were questioned whether they contained a brain or not, 40 (5.66%) when questioned if the brain was acquired in full, and in 36 (5.09%) series the annotators could not assert whether there was another body part. Annotators could not identify the sequence type for 85 (12.02%) series and the presence of contrast in 104 (14.71%) series.

In this first round of annotations, the main source of disagreements between annotations and labels was the presence of non-brain images or localisers. For example, non-brain scans had a higher rate of mistakes in sequence annotation. For further analyses, the scans labelled as ‘localiser’ in the first round of annotations, which contained under 15 slices, were ignored. If the annotation and label agreed on the absence of brain, the images were not re-annotated.

In the second round of annotations, the images with unknown sequence type (16 scans), and those with disagreement between label and annotation in which either assigned ‘T1’, ‘T2’ or ‘FLAIR’ (48 scans) were re-annotated. Additionally, a subset of images was selected out of 66 which had partial agreement between at least one sequence label and the annotation, to validate commonly occurring combination of labels which were not similar but not outright disagreeing (e.g. ‘T1’ + ‘GRE’ instead of ‘T1’ +). Scans which contained at least one mention of ‘T1’, ‘T2’ or ‘FLAIR’ in either the annotation or the label were re-annotated for contrast presence when disagreement was found or when they were annotated as ‘Unknown’. Series with ‘Unknown’ annotation for questions regarding brain presence (13), other body part presence (8) and whole brain (11) were also re-annotated. Disagreement between the annotation and the labels were also re-annotated, respectively 7, 143 and 20 scans. In case of a whole brain, the disagreement was ignored if other body parts were present and if label and annotation agreed. In total, 143 series were re-annotated for the presence of brain and other body parts, and for full brain coverage. Additionally, there were 84 images re-annotated for sequence type and contrast.

To resolve conflict between the two re-annotators, or between them and the labelling tools, 27 series were then annotated a third time. Some conflicts could not be resolved, such as 7 images having the same ‘Series Description’ (0008,103E) tag value, and thus the same label. Three of them were identified as T1 and four as T2* by the two annotators in agreement.

Between each round of annotation, the regular expressions used by the labelling tools were updated to reflect previously unknown, and to solve conflicting information and errors.

The results of the labelling tools compared to the annotation as ground truth were very good. The true positive rate ranged from 87% to 97% and the positive predictive value from 81% to 99% (Table 2). The presence of other body parts and contrast was amended to their absence, the target which we considered “positive”. This value excludes localisers for sequence type, and series without presence of brain for ‘whole brains’. The lower precision for detection of other body parts is explained by the lack of mention of any parts in the different DICOM tags, sometimes due to missing data, as well as the detection of some other head parts, such as the jaw, without mention of the brain, which often, but not always, indicate non brain scans. The lower recall for the absence of contrast is caused by the low number of studies that used intravenous (IV) contrast. During a scanning session that used IV contrast, a first image will normally be captured free of contrast, prior to the injection, however, the ‘Study Description’ (0008,1030) will indicate the presence of IV contrast nonetheless for this first series, as was commonly found.

### Permissions and governance

Our application to the PBPP, which included an industry partner and aimed to develop an AI algorithm, required 210 days spanning 4 iterations for approval from the initial submission and over 17,000 words across 33 pages.

The data flow and linkage process for the BHD framework are schematically illustrated in Figure 3. Researchers can either log into a workspace running in the NSH, over approved secure channels, where data will be provided directly alongside the tools to perform analysis. To run externally developed tools, they can build a container outside the NSH, and pull it from a public registry (such as the gihub container registry: ghcr.io). Alternatively, we plan to allow for the ADDI Workbench (https://www.alzheimersdata.org/ad-workbench) to run an analysis workflow externally, and receive the output once approved by eDRIS. It should be noted that no data leaves the EPCC TRE during this process. The TRE is divided in several zones (Figure 3). The blue zones, where eDRIS store the data, are not accessible to the researchers. The different areas the researchers have access to carry their work comprise the green zones. They will have access to subsets of the data, as defined by their project group. The zones coloured in yellow are external to the NSH and represents cloud resources, such as the ADDI workbench, or container registry.

**Figure 3.**
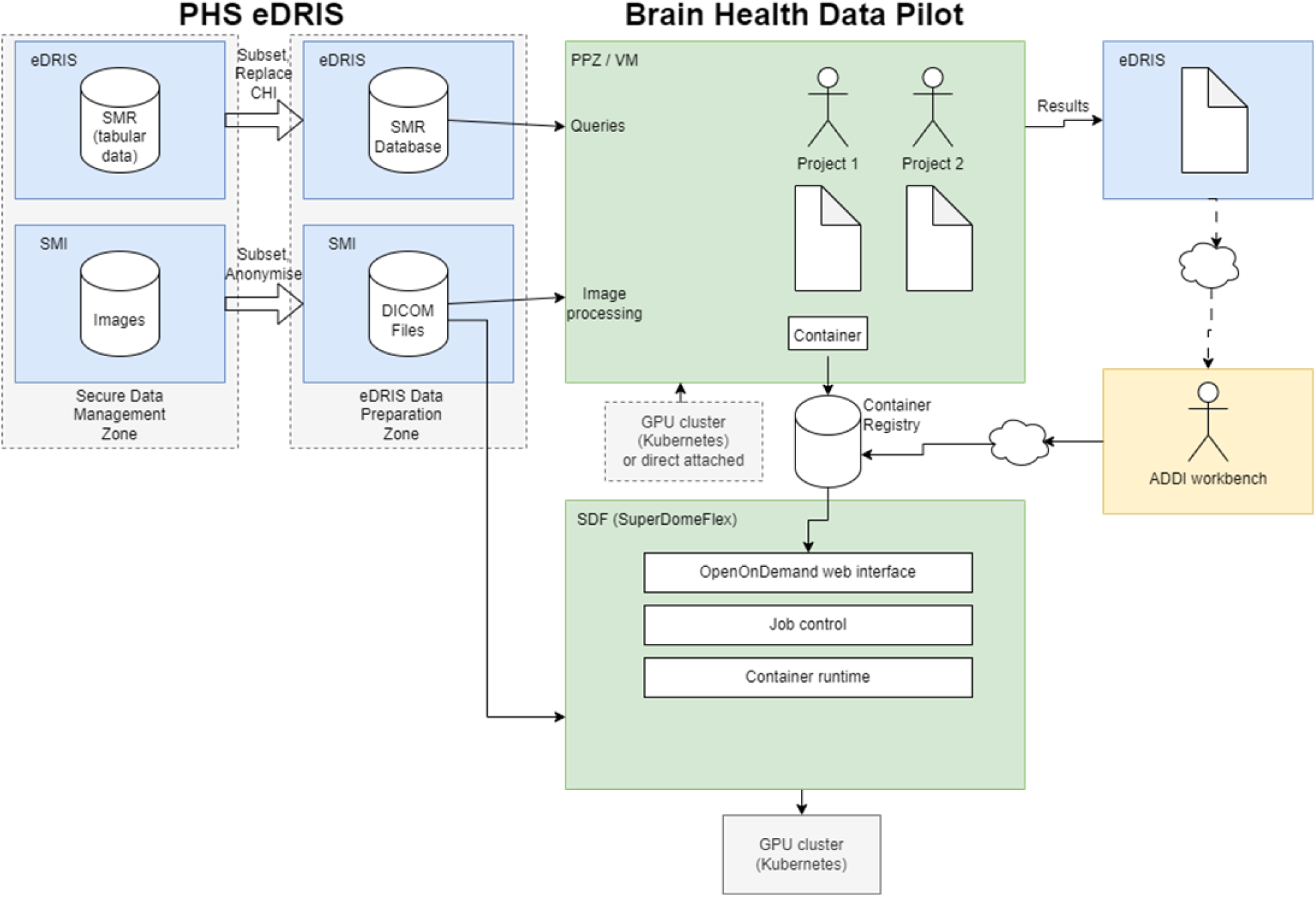
Diagram to illustrate the data flow and data linkage process in the BHD.

**Figure 4.**
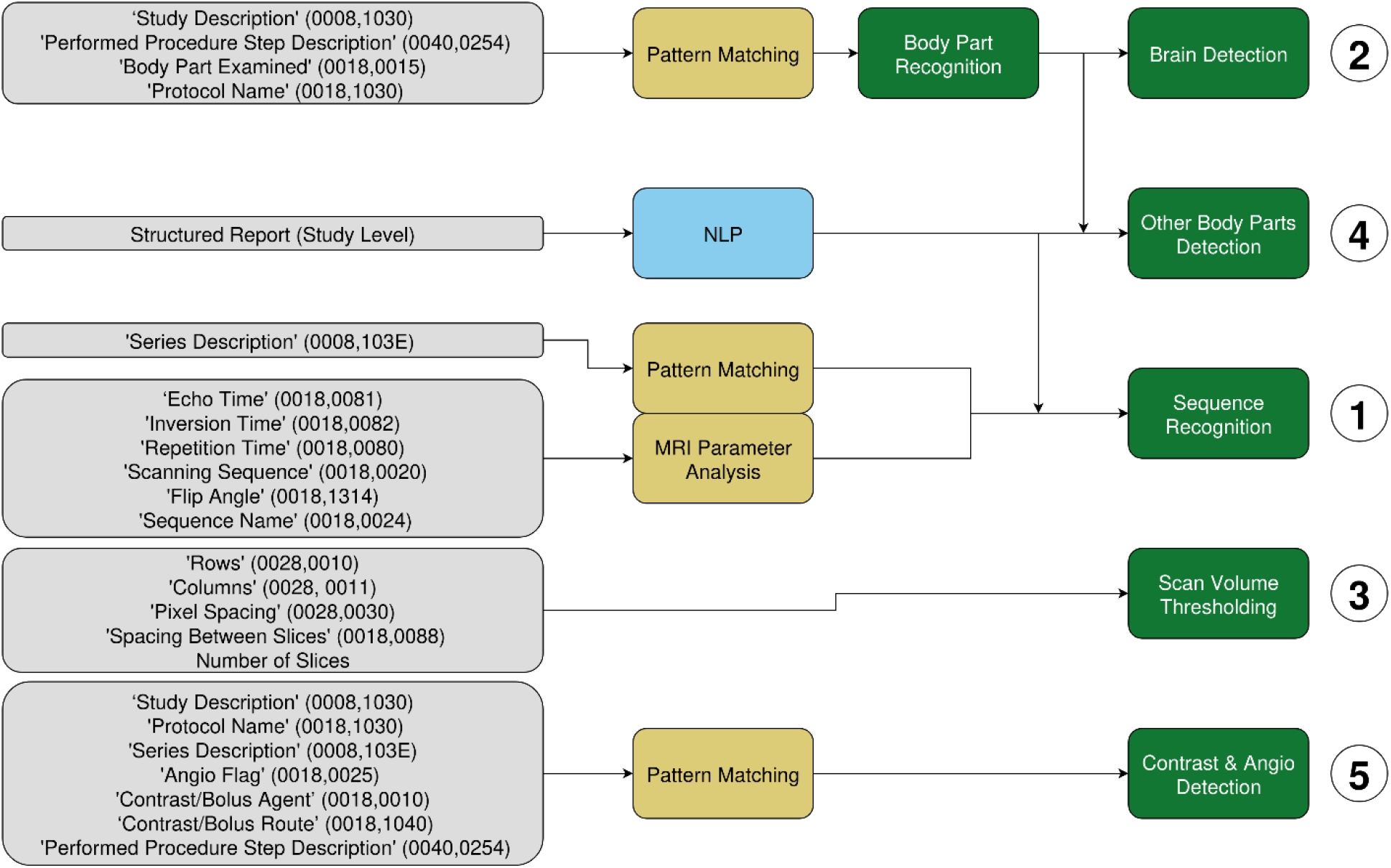
Flowchart summarizing the filtering of the MRI scans using DICOM metadata and structured report

There can be different project groups working simultaneously through the BHD. Each group may only have access to a subset of the data as necessary for their research, whether that be a subset of tables, a subset of rows, or a subset of patients. To gain an additional level of confidentiality each group will only see a CHI replaced by a pseudonymised identifier which is specific to their group. eDRIS will maintain a mapping from the Encrypted Universal Patient Identifier (EUPI) to these group-specific pseudonymised identifiers.

PHS eDRIS will prepare suitable subsets of the data for a particular research group and copy it to their working space. The eDRIS research coordinator will review any results of the researcher team to be released outside of the TRE, for example for publication.

## DISCUSSION

To our best knowledge, BHD is the first large-scale, curated brain imaging clinical dataset relevant to dementia research that is available to researchers via moderated public access. The dataset offers several advantages in addition to its large sizer: clinical relevance, long-term follow-up, co-location with a GPU cluster in a safe haven, greater population representativeness compared to many research cohorts, and accessibility for clinical researchers. The resource continues to grow in data size and computing power.

Working with health systems data presents challenges. One of them is the time elapsed for governance approvals. In our case, if governance had been applied for after the funding had been awarded, it would have represented 58% of a 1-year postdoctoral award. This would not only impact negatively on career development of the post holder but also delay the project goals. To address this barrier, BHD has co-developed a streamlined application process with PHS, which included simplified forms for pre-competitive research and faster approval timelines. Data provision was initially constrained by the limitations of the virtual machine environment, limited staff availability, and increased procedural complexity resulting in delayed access to imaging data and complicating project planning. The framework established by SCANDAN, now adopted by PHS, and the experience gained throughout the conduction of the project, are expected to accelerate data delivery for future projects. It is important however, to note that all research outputs generated within the NSH must undergo review by PHS staff prior to release.

Most imaging research is based on uniformly acquired research data. In contrast, clinical scans acquired in a routine free-at-the-point-of-service healthcare are sometimes incomplete, may be obscured by movement or other artefacts, show signs of non-relevant pathologies, may have been obtained with non-standardised protocols, and on different machines. However, such real-world data with inherent variability is essential for the development of software tools suitable for robust application in clinical practice where such heterogeneity is the norm.

Using electronic health records for dementia diagnosis has limitations. Currently, primary care data are unavailable through PHS. Hence, we relied on recorded diagnosis after an inpatient stay or death. Hospital and death records under-ascertain (false negatives) dementia in the short term and have modest reliability for dementia subtypes [15]. However, they have also previously shown high positive predictive value for all dementia diagnosis [14]. Referral reasons for scans acquisitions are not currently available, although further NLP work with reports could achieve this.

The use of head scans does raise privacy concerns due to facial recognition risks. We have mitigated these by working only in a safe haven environment, visually examining only brain slices, prohibiting facial reconstruction, limiting access to approved researchers, and having PHS strictly checking all outputs from the secure environment. Future work aims to further mitigate privacy risks by limiting the need for direct human access to data, for example by implementing software via containers. However, this work needs training of the research community, better labelling of metadata (so the data is truly FAIR), and further development of technology within the NSH environment.

There are many opportunities for further linkage to other datasets (for example community retinal imaging[16]). Such work will require further engagement with public contributors, and improvement in the security process to extract AI models trained within the NSH.

Researchers can access the BHD data by applying to eDRIS. Proposals must demonstrate a clear public benefit, and researcher-generated outputs must be added back to the dataset so every project strengthens the next. We strongly encourage cross-group collaboration. The resources available through the BHD are growing in terms of data availability, storage capacity, and computing power that are provided to researchers. We hope that this, and similar global initiatives, will ultimately contribute to improve the brain health of people worldwide.

## Data Availability

All data are available by application to eDRIS, https://publichealthscotland.scot/resources-and-tools/health-intelligence-and-data-management/electronic-data-research-and-innovation-service-edris/overview/what-is-edris/

